# Patients with COVID-19 Infection and Stroke have Higher than Expected Mortality, Regardless of the Primary Presentation

**DOI:** 10.1101/2024.03.29.24305101

**Authors:** Jintong Liu, Eric Fu, Riley Gillette, Max Wohlauer

## Abstract

**Background:** COVID-19 infection is associated with thrombotic events; however, this phenomenon is poorly understood. Few studies have reported the association between COVID-19 and stroke in the hospital setting.

**Methods:** We retrospectively reviewed and characterized all patients who presented to a single, quaternary medical center between March and December 2020 (N=603). COVID-19 positive patients who developed ischemic or hemorrhagic stroke were included in the analysis (N=66). This cohort was compared with patients who were COVID-19 negative at the time of stroke presentation in the same period (N=537). Statistical significance was evaluated using Pearson’s Chi squared test with Yates’ continuity correction and linear model ANOVA.

**Results:** Sixty-six patients had COVID-19 and Stroke. Of these patients, 22 (33.4%) patients initially presented with stroke and 44 (66.7%) initially presented with COVID-19. Patients who presented with COVID-19 and had a stroke during their hospitalization (COVID-first) had worse outcomes than patients presenting to the hospital with stroke whose COVID test became positive later in the hospitalization (stroke-first). Patients who presented with COVID-19 and had a stroke during their hospitalization had an increased rate of acute renal failure (48.9% vs 19.0%, p=0.021) and need for ventilation (60.0% vs 28.6%, p=0.017). Further, in the COVID-first cohort, the use of heparin prior to the stroke event was not associated with mortality or type of stroke (ischemic or hemorrhagic).

**Conclusion:** In the early pandemic, patients with COVID-19 infection and stroke had a higher mortality rate compared to COVID-19 negative patients with stroke. Among patients with both COVID-19 and stroke, patients presenting with COVID-19 first had worse outcomes than patients presenting with stroke first. The use of heparin prior to the stroke event was not associated with mortality or type of stroke.

## Introduction

Over 6.9 million people have died from COVID-19 worldwide and hundreds of millions more have suffered from SARS-CoV-2 associated illness. ^1^ While most symptoms involve the respiratory tract, neurologic symptoms such as altered consciousness, seizures, stroke, and sensory deficits have been extensively described. ^2^ Stroke incidence in COVID-19 patients ranges between 0.1 to 6.9% of hospitalized cases. ^3,4^

COVID-19 is associated with a hypercoagulable state and resulting thromboembolic events, including deep vein thrombosis, pulmonary embolism, and stroke. ^4–7^ Several mechanisms have been proposed including endothelial injury, inflammation, complement activation, and deactivation of antithrombotic pathways. ^8^An interesting model, based off postmortem studies and these mechanisms, postulates that injury to the lung tri-compartment (pulmonary artery, bronchial artery, and alveolus) leads to pulmonary vein thrombosis and subsequent large vessel occlusion (LVO) ^9^. In terms of demographics, COVID-19 patients who suffered strokes are also demographically different from traditional stroke patients. They are typically younger, more often without previous vascular risk factors, more likely to be male, and more likely to experience large vessel occlusions. ^3,5,10^

Despite the large number of studies on stroke and COVID-19 that have been published, a causal relationship between SARS-CoV-2 and stroke has yet to be established. ^3^ Moreover, there is a notable gap in research concerning hemorrhagic stroke within the context of SARS-CoV-2 infection, despite documented instances of hemorrhagic complications. ^4,5^ In addition to reporting occurrence and outcome, one way to explore possible correlations is by differentiating the chronologic sequence of the presenting symptoms. Our study separates stroke patient outcomes through patients’ primary presentation, whether they presented with COVID-19 or stroke first. Furthermore, we explored the relationship between timing of anticoagulation on outcomes, which has not been described previously.

## Methods

We conducted a retrospective study of 603 consecutive patients who presented with stroke in a quaternary medical center between March 2020 and December 2020. Sixty-six patients who were COVID-19 positive and 537 patients who were COVID-19 negative at the time of ischemic or hemorrhagic stroke presentation were included. Characteristics and comorbidities were identified and compared between two groups using Pearson’s Chi squared test with Yates’ continuity correction and linear model ANOVA. Missing data such as race and sex were not included in analysis. Comparisons stratified by stroke types (ischemic or hemorrhagic) and mortality rates were also performed. Within the 66 patients who were COVID-19 positive at the time of ischemic or hemorrhagic stroke presentation, we further stratified by their primary presentation, whether patients presented with COVID-19 first and developed a stroke during their hospitalization (N=45) (stroke-first) or whether stroke was the initial presenting symptom of COVID-19 (COVID-first) (N=21). Characteristics, comorbidities, and outcomes were compared between two groups. A multivariable logistic regression model was used with the presence or absence of postoperative death as the dependent variable, COVID-19 and stroke presentation, and comorbidities diabetes mellitus and chronic kidney disease (CKD) as independent variables based off significance in univariate analyses. In addition, we identified causes of death by systems for those who were COVID-19 positive at the time of stroke presentation. Lastly, for the COVID-first cohort (N=45), we distinguished those who received prophylaxis or therapeutic heparin from those who did not prior to the onset of stroke. The types of stroke and mortality rate were then compared with Pearson’s Chi-squared test with Yates’ continuity correction. Statistical significance was defined as p < 0.05. Analyses were done using R (version 4.2.0, R Foundation for Statistical Computing, Vienna, Austria).

The case report form was developed by a working group of the Vascular Surgery COVID-19 Collaborative (VASCC). ^11^ The VASCC was established on March 2, 2020, to study the impact of the ongoing COVID-19 pandemic on vascular surgical care. STROBE guidelines for observational studies were followed.

## Results

The mean age for all patients was 64.4 years old, and it was similar between both cohorts (Table 1). More patients identified as Hispanic in the COVID & Stroke cohort than patients in the Stroke Only cohort (40.9% vs 11.4%, p<0.001). The male and female distribution was similar between the two cohorts (p=0.200). Prevalence of diabetes mellitus and chronic kidney disease were higher in the COVID & Stroke cohort than in the Stroke Only cohort (p<0.001). Ischemic stroke was more prevalent than hemorrhagic stroke in all patients and in both cohorts. Mortality was significantly higher in the COVID & Stroke cohort (patients with COVID who developed a stroke) than in the Stroke Only cohort (37.9% vs 17.3%, p<0.001).

**Table 1.** Demographic Characteristics, Comorbidities, and Outcomes for all Patients. Demographic characteristics, comorbidities, and outcomes for all patients presented with stroke to a tertiary medical center between March 2020 and December 2020. COVID & Stroke cohort included patients who were COVID-19 positive at the time of stroke presentation, while Stroke Only cohort included patients who were COVID-19 negative. SD = standard deviation. p values compared COVID & Stroke cohort and Stroke Only cohort.

| **Table 1. Demographic Characteristics, Comorbidities, and Outcomes for all Patients** |  |  |  |  |
| --- | --- | --- | --- | --- |
| **Variables** | **All Patients**<br>**(N=603)** | **COVID &**<br>**Stroke (N=66)** | **Stroke Only**<br>**(N=537)** | **p values** |
| Mean Age (SD) | 64.40 (17.90) | 62.11 (14.97) | 64.68 (18.22) | 0.270 |
| Race |  |  |  | <0.001 |
| Native American | 4 (0.7%) | 0 (0.0%) | 4 (0.7%) |  |
| Asian or Pacific Islander | 24 (4.0%) | 5 (7.6%) | 19 (3.5%) |  |
| Black, Non-Hispanic | 98 (16.3%) | 4 (6.1%) | 94 (17.5%) |  |
| Hispanic | 88 (14.6%) | 27 (40.9%) | 61 (11.4%) |  |
| White, Non-Hispanic | 290 (48.1%) | 26 (39.4%) | 264 (49.2%) |  |
| Unknown/Other | 99 (16.4%) | 4 (6.1%) | 95 (17.7%) |  |
| Sex |  |  |  | 0.200 |
| Female | 240 (39.8%) | 25 (37.9%) | 215 (40.0%) |  |
| Male | 282 (46.8%) | 41 (62.1%) | 241 (44.9%) |  |
| Unknown/Other | 81 (13.4%) | 0 (0.0%) | 81 (15.1%) |  |
| Comorbidities |  |  |  |  |
| Diabetes Mellitus | 182 (30.2%) | 37 (56.1%) | 145 (27.0%) | <0.001 |
| Coronary Artery Disease | 66 (10.9%) | 6 (9.1%) | 60 (11.2%) | 0.762 |
| Congestive Heart Failure | 43 (7.1%) | 7 (10.6%) | 36 (6.7%) | 0.363 |
| History of Stroke | 112 (18.6%) | 11 (16.7%) | 101 (18.8%) | 0.799 |
| Transient Ischemic Attack | 25 (4.1%) | 6 (9.1%) | 19 (3.5%) | 0.071 |
| Hyperlipidemia | 184 (30.5%) | 24 (36.4%) | 160 (29.8%) | 0.341 |
| Hypertension | 371 (61.5%) | 45 (68.2%) | 326 (60.7%) | 0.297 |
| Atrial Fibrillation | 57 (9.5%) | 5 (7.6%) | 52 (9.7%) | 0.742 |
| Chronic Kidney Disease | 37 (6.1%) | 13 (19.7%) | 24 (4.5%) | <0.001 |
| Obesity (BMI > 30 kg/m<sup>2</sup>) | 21 (3.5%) | 3 (4.5%) | 18 (3.4%) | 0.886 |
| Tobacco Use | 130 (21.6%) | 10 (15.2%) | 120 (22.3%) | 0.237 |
| Stroke Type |  |  |  | 0.331 |
| Hemorrhagic | 166 (27.5%) | 22 (33.3%) | 144 (26.8%) |  |
| Ischemic | 437 (72.5%) | 44 (66.7%) | 393 (73.2%) |  |
| Death | 118 (19.6%) | 25 (37.9%) | 93 (17.3%) | <0.001 |

Stratified based on the stroke type (hemorrhagic or ischemic), similar results were found (Supplemental Tables 1 and 2). However, for patients presented with hemorrhagic stroke, the mortality rates were not statistically significant between the COVID & Stroke cohort and the Stroke Only cohort (40.9% vs 26.4%, p=0.249). When modeled in a multivariable logistic regression framework, Stroke and COVID-19 presentation (OR: 0.41, 95% C.I. (0.23, 0.73), p=0.002) and CKD (2.36, (1.12-4.82), p = 0.02) were significantly associated with postoperative death, diabetes mellitus did not have a significant association (1.15, (0.73,1.79), p=0.528).

Patients who presented with stroke only were less likely to experience postoperative death compared to those with both stroke and COVID-19. Patients with CKD were more likely to experience postoperative death compared to non-CKD patients. In terms of revascularization, out of 44 patients with ischemic stroke and COVID-19, only one patient received mechanical thrombectomy and only three patients received IV thrombolysis (2.3% and 6.8% respectively).

When stratified by primary presentation (COVID or Stroke) within the COVID & Stroke cohort, demographic characteristics and comorbidities were not significantly different between the two groups (Table 2). Type of strokes and mortality rates were also not significantly different between two groups. Patients with COVID-19 as the primary presentation were associated with worse outcomes including increased acute renal failure and need for ventilation rates when compared to patients presented with stroke first (48.9% versus 19.0%, p=0.021 and 60.0% versus 28.6%, p=0.017, respectively).

**Table 2.** Demographic Characteristics, Comorbidities, and Outcomes for Patients with COVID-19 and Stroke. Demographic characteristics, comorbidities, and outcomes for patients who were COVID-19 positive when presented with stroke to a tertiary medical center between March 2020 and December 2020. COVID-first group included patients who were diagnosed with COVID-19 prior to stroke presentation, while Stroke-first group included patients who were diagnosed with COVID-19 after stroke presentation. SD = standard deviation.

| **Table 2. Demographic Characteristics, Comorbidities, and Outcomes for Patients with COVID-19 and Stroke** |  |  |  |
| --- | --- | --- | --- |
| **Variables** | **COVID-First (N=45)** | **Stroke-First (N=21)** | **p values** |
| Mean Age (SD) | 62.73 (12.51) | 60.76 (19.52) | 0.622 |
| Race |  |  | 0.165 |
| Native American | 0 (0.0%) | 0 (0.0%) |  |
| Asian or Pacific Islander | 3 (6.7%) | 2 (9.5%) |  |
| Black, Non-Hispanic | 2 (4.4%) | 2 (9.5%) |  |
| Hispanic | 22 (48.9%) | 5 (23.8%) |  |
| White, Non-Hispanic | 17 (37.8%) | 9 (42.9%) |  |
| Unknown/Other | 1 (2.2%) | 3 (14.3%) |  |
| Sex |  |  | 0.166 |
| Female | 14 (31.1%) | 11 (52.4%) |  |
| Male | 31 (68.9%) | 10 (47.6%) |  |
| Comorbidities |  |  |  |
| Diabetes Mellitus | 27 (60.0%) | 10 (47.6%) | 0.498 |
| Coronary Artery Disease | 2 (4.4%) | 4 (19.0%) | 0.144 |
| Congestive Heart Failure | 6 (13.3%) | 1 (4.8%) | 0.532 |
| History of Stroke | 7 (15.6%) | 4 (19.0%) | 1.000 |
| Transient Ischemic Attack | 3 (6.7%) | 3 (14.3%) | 0.587 |
| Hyperlipidemia | 15 (33.3%) | 9 (42.9%) | 0.635 |
| Hypertension | 29 (64.4%) | 16 (76.2%) | 0.503 |
| Atrial Fibrillation | 3 (6.7%) | 2 (9.5%) | 1.000 |
| Chronic Kidney Disease | 9 (20.0%) | 4 (19.0%) | 1.000 |
| Obesity (BMI > 30 kg/m<sup>2</sup>) | 3 (6.7%) | 0 (0.0%) | 0.564 |
| Tobacco Use | 8 (17.8%) | 2 (9.5%) | 0.615 |
| Days Between COVID and Stroke | 19.30 (16.73) | 1.00 (1.76) |  |
| Stroke Type |  |  | 0.400 |
| Hemorrhagic | 17 (37.8%) | 5 (23.8%) |  |
| Ischemic | 28 (62.2%) | 16 (76.2%) |  |
| Outcomes |  |  |  |
| Acute Renal Failure | 22 (48.9%) | 4 (19.0%) | 0.021 |
| Ventilation | 27 (60.0%) | 6 (28.6%) | 0.017 |
| Tracheostomy | 5 (11.1%) | 0 (0.0%) | 0.112 |
| ECMO | 4 (8.9%) | 0 (0.0%) | 0.159 |
| Myocardial infarction | 5 (11.1%) | 0 (0.0%) | 0.112 |
| Death | 18 (40.0%) | 7 (33.3%) | 0.804 |

We also characterized causes of death in patients who were COVID-19 positive at the time of stroke presentation (the COVID & Stroke cohort) (Table 3). Among all patients, neurological causes of death were most common, followed by pulmonary causes.

**Table 3.** Causes of Death by Systems in the COVID & Stroke Cohort. Causes of death by system in patients who were COVID-19 positive at the time of stroke presentation.

| **Table 3. Causes of Death by Systems in the COVID & Stroke Cohort** |  |  |  |
| --- | --- | --- | --- |
|  | **COVID-First** | **Stroke-First** | **Total** |
| Pulmonary | 7 | 2 | 9 |
| Neurological | 7 | 4 | 11 |
| Multi-organs | 3 | 0 | 3 |
| Unknown | 1 | 1 | 2 |

In the COVID-first cohort (N=45), the use of heparin prior to the stroke event was not associated with type of stroke or mortality (Table 4). Among the 29 patients who received heparin, 13 developed hemorrhagic stroke, and 16 developed ischemic stroke, which was not significantly different from those who did not receive heparin (p=0.321). Mortality rate was 41.4% in those who received heparin while 37.5% in those who did not receive heparin (p=1.000).

**Table 4.** Stroke Types and Mortality for Patients Received Heparin or Not Prior to Stroke Onset. Stroke types and mortality for patients presented diagnosed with COVID-19 prior to stroke presentation to a tertiary medical center between March 2020 and December 2020, stratified by whether received heparin or not prior to stroke onset.

| **Table 4. Stroke Types and Mortality for Patients Received Heparin or Not Prior to Stroke Onset** |  |  |  |
| --- | --- | --- | --- |
|  | **Received Heparin** | **Did not Receive Heparin** | **p values** |
| Total | 29 | 16 |  |
| Stroke Type |  |  | 0.321 |
| Hemorrhagic | 13 (44%) | 4 (25%) |  |
| Ischemic | 16 (55.2%) | 12 (75%) |  |
| Death | 12 (41.4%) | 6 (37.5) | 1.000 |

## Discussion

In this retrospective cohort study, we found significant differences in the demographics of the cohort who had a stroke versus those who had both a stroke and COVID-19 in terms of the presence of diabetes, CKD, and race. We also found a significant difference in mortality and morbidity. Additionally, we looked specifically at those with hemorrhagic stroke, as opposed to ischemic stroke, and did not find a correlation with heparin use in our cohort in contrast to previous studies.

### Stroke Overall

In terms of both ischemic and hemorrhagic stroke, our key findings include a greater prevalence of diabetes, CKD, and Hispanic ethnicity in those with COVID-19 infection and stroke than those with only a stroke, higher mortality in those with both, and worse outcomes for those presented with COVID-19 first then stroke as opposed to stroke first then COVID-19.

It has been widely observed that diabetes and CKD are both risk factors associated with increased COVID-19 infection rates and the severity. ^12,13^ For our cohort, diabetes mellitus (27% vs 56%, p<0.001) and CKD (4.5% vs 20%, p<0.001) were both significantly more prevalent in those who had COVID and stroke than those who presented only with stroke. Not only are patients with these comorbidities more likely to be infected by SARS-CoV2, but they could also be more likely to suffer stroke while infected.

Additionally, there is a significant difference in the racial makeup of those who suffered only a stroke (n=537) versus those who had both COVID-19 and stroke (n=66, p<0.001). This contrasts with no significant difference found between those who had COVID-19 first then stroke versus having stroke first then COVID-19. This significant difference aligns with general COVID-19 trends where the infection disproportionally affects those in the minority. ^14^ We observed that those who identify as Hispanic were a significantly greater percentage of the overall COVID-19 and stroke patient population than stroke only (41% vs 13%).

In terms of mortality, our findings revealed that those with COVID-19 and stroke during the same hospital stay exhibited a higher death rate than those who suffered from a stroke only (38% vs 17%, p<0.001) in the early stages of pandemic. This builds upon earlier research indicating elevated mortality in COVID-19 patients with stroke compared to those without, ^15^ as well as an increased risk of severe disability and mortality when compared to propensity-matched non-COVID-19 stroke patients. ^16,17^ Our clinical observations align with this conclusion, suggesting that a higher disease burden correlates with increased mortality, particularly in diseases with inherently high mortality rates. Furthermore, even within the same disease, more severe infections were associated with a higher incidence of stroke. ^15^

Interestingly, patients who presented with COVID-19 and had a stroke during their hospitalization (COVID-first) had worse outcomes than patients presenting to the hospital with stroke whose COVID-19 test became positive later in the hospitalization (stroke first). Those who were admitted with symptoms of COVID-19 and endured a stroke during their hospitalization were more likely to have acute renal failure (49% vs 19%, p=0.021) and need ventilation (60% vs 29%, p=0.017). This could reflect the differences in the severity of the COVID-19 infection in the two groups. For patients with stroke first in this study, they were more likely to have an asymptomatic COVID-19 infection that was not severe enough to have been tested. For those with COVID-19 first, they were likely hospitalized with COVID-19 pneumonia and needed ventilation and were also more likely to experience acute renal failure, a common morbidity of COVID-19. ^18^

### Hemorrhagic Stroke

Despite COVID-19 being mostly characterized as a hypercoagulable state, bleeding complications such as increased morbidity/mortality post-stroke revascularization and hemorrhagic strokes have been reported. ^4,6,19^ Previous studies found that while inherent risk of hemorrhagic stroke low, overall risk is increased in the setting of full-dose anticoagulation use for thromboembolism prevention as part of COVID-19 management. ^20^ Thus, our study strived to delineate between ischemic and hemorrhagic stroke, separate outcome by heparin use, and compare comorbidities within each type of stroke between those who had stroke only versus those who had both COVID-19 and stroke.

Among patients with hemorrhagic stroke, hyperlipidemia was more likely in those who had both stroke and COVID-19 than those who only had a stroke (p=0.046). While previous studies found that hyperlipidemia is not significantly associated with all-cause mortality in COVID-19 patients, it in combination with COVID-19 may specifically predispose patients to hemorrhagic stroke. ^21^ Second, in contrast with previous studies, we did not find hemorrhagic stroke to be more common in patients with pre-existing hypertension and diabetes mellitus prior to COVID-19 infection. ^22^ Third, in the COVID-first cohort (N=45), the use of heparin prior to the stroke event showed no significant correlation with mortality (p=1.000) or type of stroke (p=0.321). This outcome is surprising given anticoagulation is a known risk of hemorrhage. ^23^ When compared to findings from a previous a multinational study, our research revealed a higher prevalence of hemorrhagic strokes (33.3% versus 21.1%).^24^ Contrary to expectations, our results suggest that the use of heparin prior to a stroke does not seem to contribute to the elevated incidence of hemorrhagic strokes. This intriguing observation prompts the need for further exploration and investigation.

Hemorrhagic stroke in patients with COVID-19 has not been heavily studied and mechanisms are still being elucidated. In combining our findings with those of others, several possible mechanisms begin to emerge. COVID-19 disproportionately affects those with underlying hypertension and diabetes and further elevates blood pressure through ACE2 dysregulation and endothelial dysfunction. ^23,25^ Furthermore, anticoagulant use has become wide-spread in COVID-19 treatment. ^23,25^ Dysregulated fibrinolysis could be another co-existing or separate mechanism.^20^ As such, patient risk factors should be considered when starting anticoagulation therapy. ^20,25^

### Limitations

Our study is limited by data from a single center and the retrospective nature of the study. The study is a snapshot in time, focusing on the 66 patients with both COVID-19 and stroke who presented from during the initial waves of COVID-19 (March to December of 2020) and was prior to the introduction of vaccines. This may limit the generalizability of the study. Second, our examination on heparin-use and its resultant morbidity and mortality was done retrospectively.

Additionally, our ability to conduct statistical analyses was limited due to the small number of patients undergoing revascularization. Only one out of the 44 patients with ischemic stroke and COVID-19 underwent mechanical thrombectomy, while just three received IV thrombolysis (2.3% and 6.8%, respectively). These figures are notably lower when compared to a previous multinational study, where 7.4% of patients underwent mechanical thrombectomy and 13.6% received IV thrombolysis. ^24^ The majority of patients in our study faced challenges in receiving revascularization, either due to their unstable condition, being outside the optimal revascularization window, or lacking indications for the procedure. Despite the absence of formal analysis in our study, a recent international multicenter retrospective cohort study suggested that patients with COVID-19 who underwent revascularization experienced worse outcomes than their counterparts without COVID-19 who underwent similar interventions. ^19^

In summary, our study retrospectively examined patients at a tertiary medical center during the initial wave of COVID-19 and found several previously less reported trends among stroke patients. First, patients with COVID-19 infection and stroke had a higher mortality rate than patients with only stroke. Second, among patients with both COVID-19 and stroke, patients presenting with COVID-19 first had worse outcomes, including higher frequency of acute renal failure and need for ventilation, than patients presenting with stroke first. Third, contrary to previous studies, the use of heparin prior to the stroke event was not associated with mortality or type of stroke.

## Data Availability

Dr. Wohlauer had full access to all the data in the study and takes responsibility for its integrity and the data analysis.

## Acknowledgments

We would like to thank the Medical Student Summer Research Program (MSRP) and Department of Surgery at the University of Colorado School of Medicine for providing time and support to perform this project. We would like to thank the Division of Neurology at the University of Colorado School of Medicine for their collaboration, and for providing valuable input and expertise.

## Non-standard Abbreviations and Acronyms

(CKD): chronic kidney disease
(ACE2): angiotensin-converting enzyme 2

## Supplemental Materials

Tables S1-S2

